# Chromosomal microarray analyses from 5,778 patients with neurodevelopmental disorders and congenital anomalies in Brazil

**DOI:** 10.1101/2022.03.08.22272093

**Authors:** Ana C. V. Krepischi, Darine Villela, Silvia Souza da Costa, Patricia C. Mazzonetto, Michele P. Migliavacca, Fernanda Milanezi, Juliana G. Santos, Rodrigo Guarischi-Sousa, Gustavo Campana, Fernando Kok, David Schlesinger, Joao Paulo Kitajima, Francine Campagnari, Angela M. Vianna-Morgante, Peter L. Pearson, Carla Rosenberg

## Abstract

Chromosomal microarray analysis (CMA) has been recommended and practiced routinely since 2010 in USA and Europe as the first-tier cytogenetic test for patients with unexplained neurodevelopmental delay/intellectual disability, autism spectrum disorders, and/or multiple congenital anomalies. However, in Brazil, the use of CMA is still limited, due to its high cost and complexity of the combination of private and public health systems. Although the country has one of the world largest single payer public healthcare system, nearly all patients referred for CMA come from the private sector. This reflects on the small number of CMA studies in Brazilian cohorts. This study is by far the largest Brazilian cohort (n=5,788) studied by CMA and results from a joint collaboration formed by the University of Sao Paulo and three private genetic diagnostic centers to investigate the genetic bases of neurodevelopmental disorders and congenital abnormalities. It is common practice to investigate the inheritance of VUS; however, our results indicate an extremely low cost-benefit of this approach, and strongly suggest that in cases of limited budget, investigation of the parents of VUS carriers using CMA should not be prioritized. Another aspect discussed is the classification of variants of low penetrance, once CNV classification is mostly designed to Mendelian or highly-penetrant variants.

## INTRODUCTION

Chromosomal microarray analysis (CMA), including array-comparative genomic hybridization (aCGH) and SNP-array, has become the gold standard procedure to detect copy number variations (CNVs) in the clinical setting. Because CMA offers a much higher diagnostic yield (15-20%) than the conventional G-banded karyotype (∼3%), the test is recommended as the first-tier cytogenetic test for patients with unexplained neurodevelopmental delay/intellectual disability, autism spectrum disorders, or multiple congenital anomalies^1^. It is noteworthy that G-banded karyotype should be offered only for patients with obvious chromosomal syndromes (e.g., Down syndrome), a family history of chromosomal rearrangement, or a history of multiple miscarriages. However, in Brazil, detection of chromosomal alterations is still performed mainly by karyotype, due to the high taxation costs of imported microarray material, and relatively cheap technical labour. As a result, the number of CMA studies in cohorts of patients with neurodevelopmental disorders and congenital anomalies is very scarce, and their sample size is typically small (< 500 individuals)^2–6^. Nonetheless, the few previous investigations reported a diagnostic rate that ranges from 15 to 22%, similar to what is cited in the literature^7–12^.

The clinical interpretation of CMA results sometimes can be challenging. Although it is now possible to screen the human genome for CNVs at a high resolution, contributing to the identification of several clinically recognizable syndromes, there are a large number of variants that are rare and unique to a particular individual or family. While some of them can be confidently predicted to be either pathogenic or benign, in many cases, pieces of evidence are missing, leaving us with many variants of uncertain significance (VUS)^13^. Hence, variant classification in often complicated; usually the criteria applied in the interpretation of a CNV include inheritance, size, type (duplication or deletion), and gene content, with support of multiple database resources for annotation^14,15^. Considering that CMA data from the Brazilian population is underrepresented in the literature and public databases, this study was established to provide a collection of genomic data from 5,778 patients with various neurodevelopmental disorders, all genotyped using a high-resolution SNP-array platform. This is by far the largest Brazilian cohort investigated in the diagnostic CMA routine; by creating this data resource we aimed to leverage an overview all cytogenetic alterations found in a clinical CMA, and document the CNVs that are clinically relevant to the diagnosis of neurodevelopmental disorders.

## MATERIAL AND METHODS

### Casuistic

The cohort presented here is the result of a joint collaboration including the Human Genome and Stem Cell Research Center of the Institute of Biosciences, University of São Paulo (IB-USP), and three private diagnostic centers located in the state of São Paulo (DASA, Mendelics, and Deoxi Biotechnology [extinct center]) to provide the largest copy number data from patients investigated in a clinical CMA routine in Brazil. Despite the diagnostic centers being in the same state in Brazil, patients were referred from different regions of the country. A total of 5,778 patients children underwent CMA, between 2010 and 2020, for presenting a general neurodevelopmental disorder and/or congenital abnormalities without an evident cause. This study was approved by the IB-USP Research Ethical Committee, and informed consent was obtained from the patients’ parents or guardians.

### Chromosomal microarray analysis (CMA) - SNP-array

Genomic DNA samples were extracted from peripheral blood cells or saliva following standard procedures. SNP-array experiments were performed using the Illumina Infinium CytoSNP 850K BeadChip (Illumina, San Diego, USA), except for 810 cases which were carried out using the Affymetrix CytoScan 750K Array (Affymetrix, Santa Clara, USA). Data were analyzed using either the BlueFuse™ Multi Analysis (Illumina, San Diego, USA) or the Chromosome Analysis Suite - ChAS Software (Affymetrix, Santa Clara, USA). Log2 ratio and B Allele Frequency (BAF) values were plotted along chromosomal coordinates, allowing the detection of both copy number changes and copy neutral regions of homozygosity (ROH).

### Variant analysis and clinical interpretation

Copy number variants were classified for their clinical impact according to the *American College of Medical Genetics* (ACMG) guidelines^15^. The criteria for variant classification were as follow:

- Pathogenic = when the CNVs (1) were more than 4 Mb in length harboring genes; (2) overlapped with regions associated with OMIM morbid genes or DECIPHER/ClinGen microdeletion/microduplication syndromes; (3) deleted haploinsufficiency OMIM genes;
- Likely Pathogenic = when the CNVs (1) were deletions partially affecting haploinsufficiency OMIM genes; (2) were 1-4 Mb in length harboring genes;
- VUS = when the CNVs (1) were duplications containing MIM genes; (2) were deletions, containing recessive MIM genes; or (3) when the segment was larger than 300 kb and harbor genes.

The common variants, i.e., those commonly reported in curated databased (DGV) were disregarded from this study. Chromosomal rearrangements were defined by the presence of more than one large CNV in different or in the same chromosome (e.g.: chromosomes derived from translocations and inversions). In particular, copy neutral ROHs restricted to a single chromosome, known to harbor imprinted regions, were considered pathogenic and likely representing uniparental disomy (UPD). ROH >10 Mb or at least two ROH >5 Mb were considered indicative of consanguinity, albeit not Pathogenic *per se*.

## RESULTS

### Diagnostic rate of the cohort

An overview of the results obtained in this study is shown in **Figure 1**. Out of the 5,778 patients with neurodevelopmental disorders or congenital abnormalities investigated, relevant CNVs were detected in 2,031 individuals, and were classified in three main categories: (i) Pathogenic; (ii) Likely Pathogenic and (iii) Variants of Unknown Significance (VUS); the corresponding frequency of each of these categories were 52% 5% and 43%, respectively (**Figure 2A**). Taking into account just the Pathogenic and Likely Pathogenic variants, the overall diagnostic yield in our cohort was 20%. .As expected, Pathogenic CNVs accounted for the largest proportion of diagnostic alterations, and are divided into seven clinically relevant classes of variants, as presented in **Figure 2B**.

**Figure 1.**
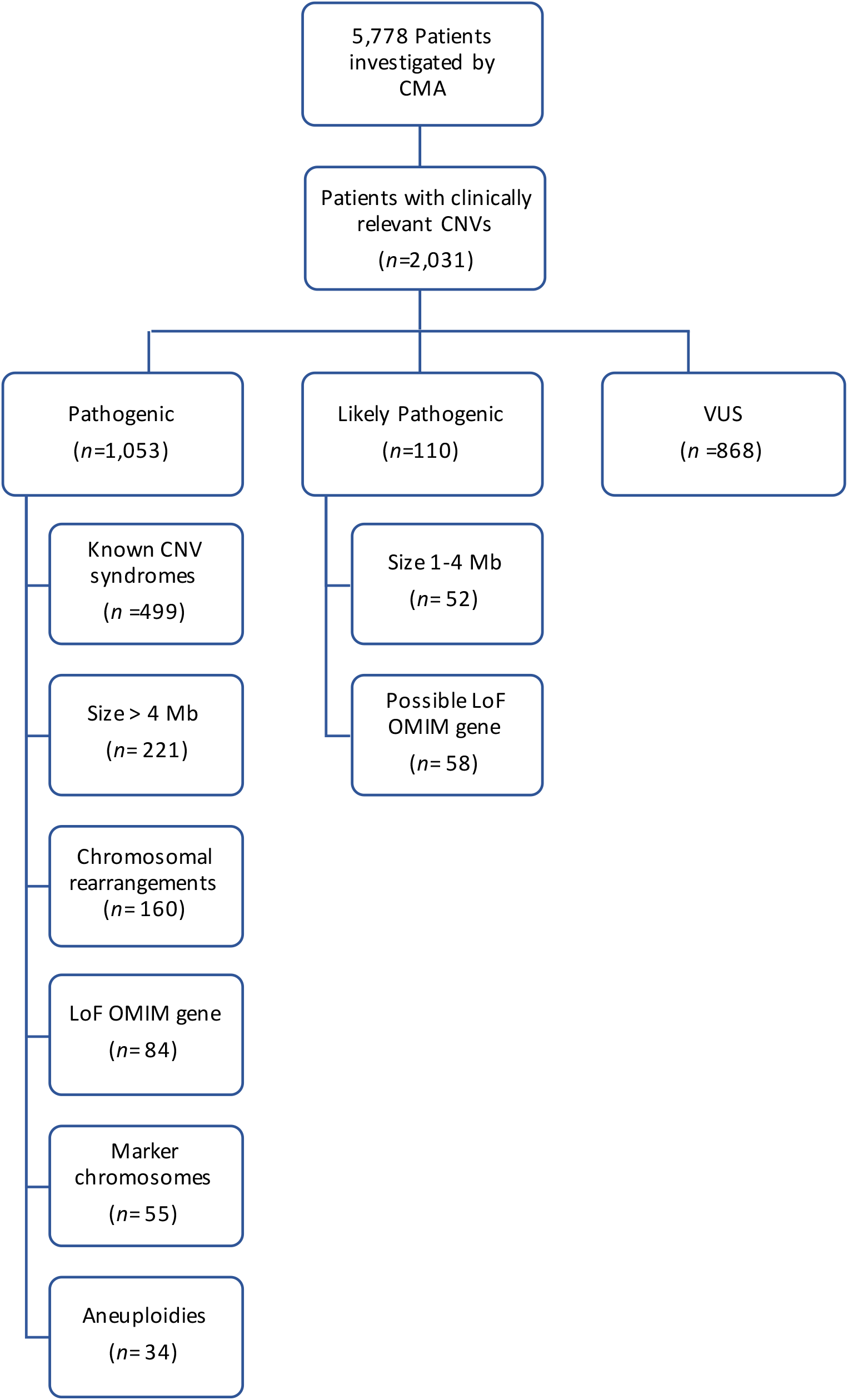
An overview of all relevant copy number variations (CNVs) identified in this study. The figure shows that, from a total of 5,778 patients with neurodevelopmental disorders were referred for chromosomal microarray analysis (CMA), 2,031 presented potentially relevant variants, classified into three main categories: (i) pathogenic CNVs; (ii) likely pathogenic CNVs; and (iii) variants of unknown significance (VUS). The total number of cases corresponding to each category is presented in the diagram.

**Figure 2.**
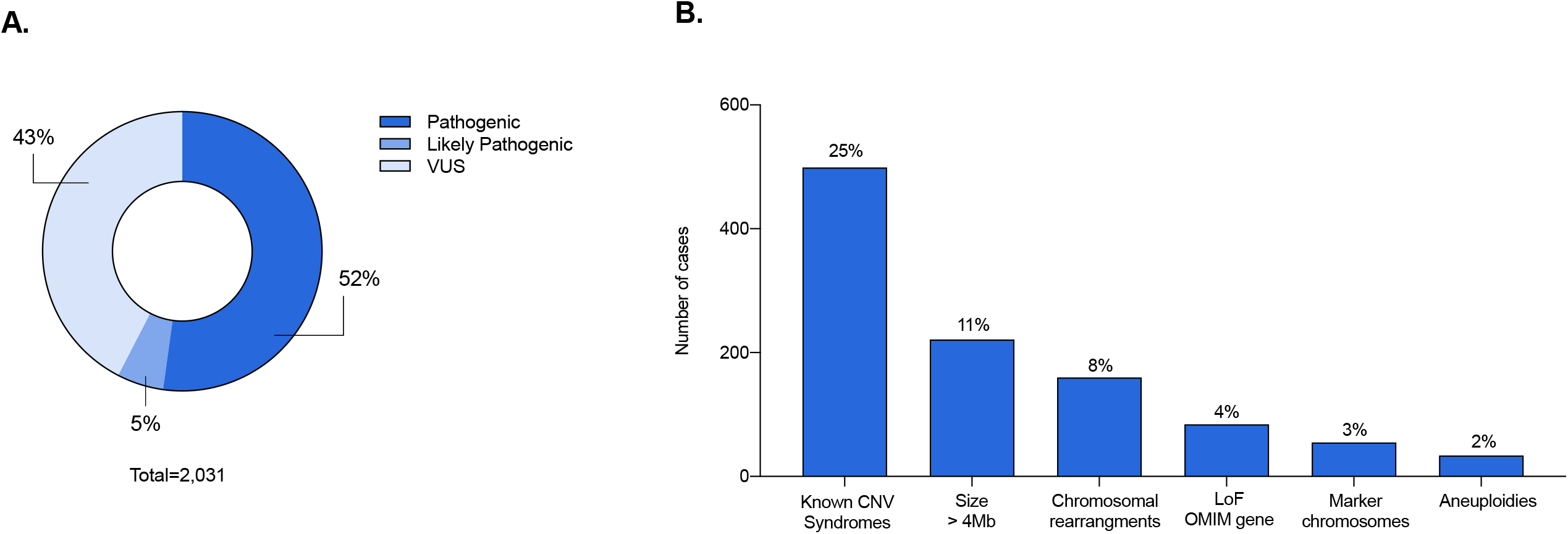
Diagnostic rate of the cohort. (A) It is shown the frequency of each of the three main CNV categories: (i) Pathogenic; (ii) Likely Pathogenic and (iii) Variants of Unknown Significance (VUS) (52% 5% and 42%, respectively). Considering just the Pathogenic and Likely Pathogenic CNVs, the overall diagnostic yield was 20% (1,178/5,778). (B) Distribution of the pathogenic CNVs, which account for the largest proportion of diagnostic alterations, displayed by frequency order: (i) known CNV syndromes (47%), (ii) CNV > 4Mb (21%), (iii) complex rearrangements (15%), (iv) loss-of-function (LoF) MIM gene (8%), (v) marker chromosomes (5%), (vi) aneuploidies (3%).

### Aneuploidies and marker chromosomes

Sex chromosome (SCA) and autosomal aneuploidies accounted for 34 cases (2%) in our cohort. Considering them as a separate groups, the proportion was very similar between SCA and autosomal trisomies: 16 and 18 cases, respectively. SCA comprise 47, XXX, 48,XXYY, 47,XYY, 47,XXY, and 45,X; the most frequent being 47,XXY (Klinefelter syndrome), found in 8/34 cases (24%). Among the autosomal trisomies, the most common was trisomy 21, found in 12/34 (35%), followed by trisomy 13 in 2/34 (6%). Excluding the known viable autosomal trisomies, an extra copy of other autosomes was only observed in mosaics, as it was the case with chromosomes 8, 9, 14 and 22. The frequency of each aneuploidy is shown in **Figure 3A**. Marker chromosomes were identified in 55 patients (3%). Marker chromosome 15 was the most frequent, corresponding to 20/55 (36%). Other markers originated from chromosomes 7, 8, 9. 10, 11, 12, 13, 18, 19, 22, X and Y; except for those derived from the sex chromosomes, all were supernumerary marker chromosomes (**Figure 3B)**.

**Figure 3.**
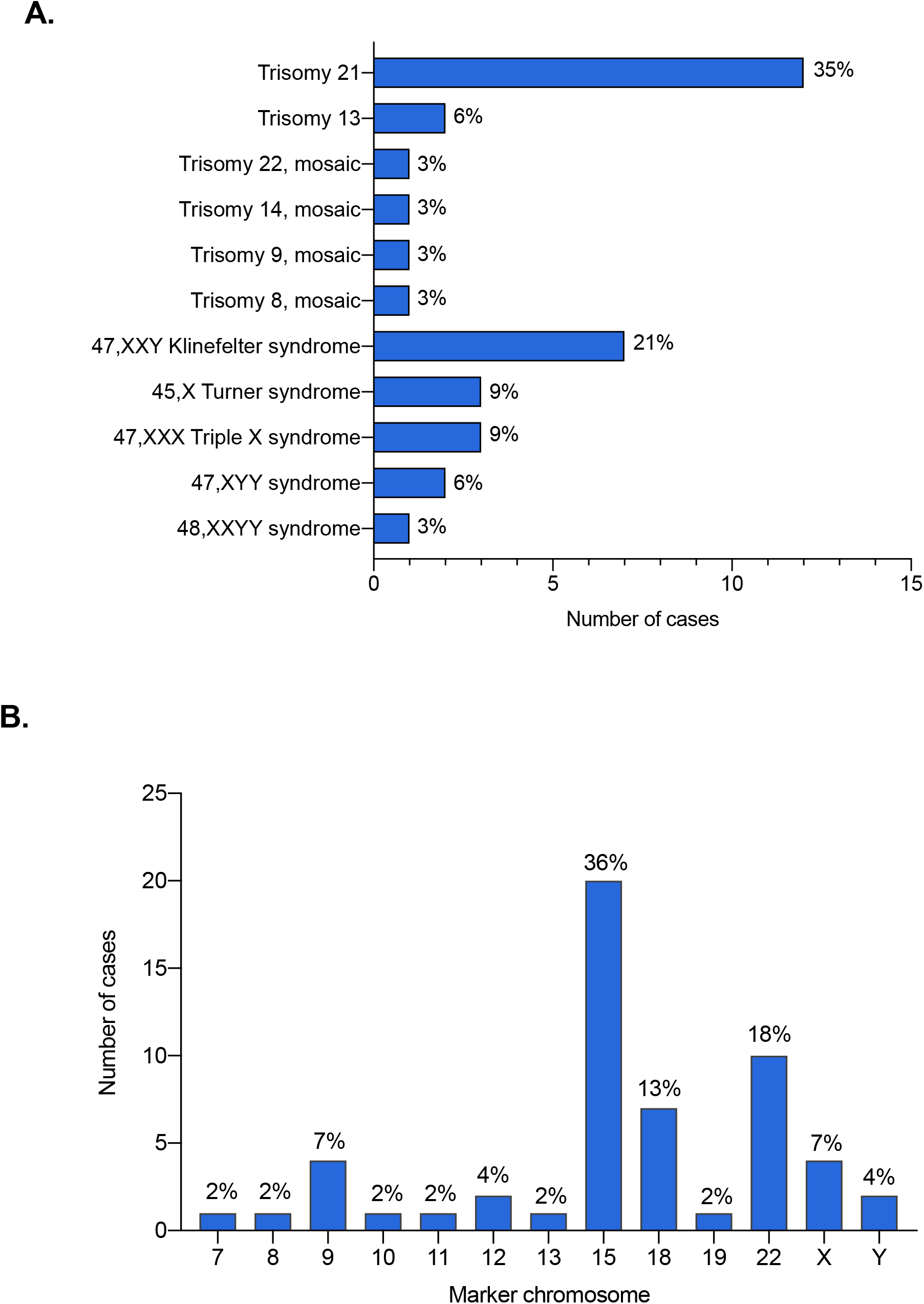
Frequency of aneuploidies and marker chromosomes. (A) Sex chromosome aneuploidies (SCA) and autosomal trisomies accounted for a total of 34 cases (2%), in which 16 correspond to SCA and 18 to autosomal trisomies. The histogram shows the frequency of aneuploidies for each chromosome. (B) Marker chromosomes were found in 55 patients (3%). The frequency of marker chromosomes according to its origin.

### Structural copy number variants

Within the Pathogenic CNV category, a total of 499 patients presented alterations associated to microdeletion and microduplication syndromes (48 deletions and 23 duplications). The five most frequent syndromes were 22q11.2 deletion (73/499 (14,6%), MIM#188400); 15q13.3 reciprocal duplication encompassing only the *CHRNA7* gene^16^ (33/499 (6,6%)); 16p11.2 deletion (25/499 (5%) MIM#611913); 15q11.2 deletion (24/499 (4,8%) MIM#615656); and Prader-Willi/Angelman syndrome (23/499 (4,6%) MIM#176270/105830, respectively). A detailed listing and frequency of each of the 71 clinical syndromes are shown in **Table 1** and **Figure 4**. Importantly, among those syndromes, there were 13 known to confer susceptibility to neurodevelopmental disorders or, in other words, present reduced penetrance; such susceptibility CNVs were detected in 144 patients, 38 of whom carried an additional variant, observed only in autosomes and mainly classified as VUS (**Figure 5**). In addition to the CNV syndromes, we detected large CNVs (> 4Mb) in 221/964 cases (23%). Chromosomal rearrangements, and loss-of-function (LoF) mutations in haploinsufficiency MIM genes accounted for 160 and 84 cases, respectively (16% and 9%).

**Table 1.**
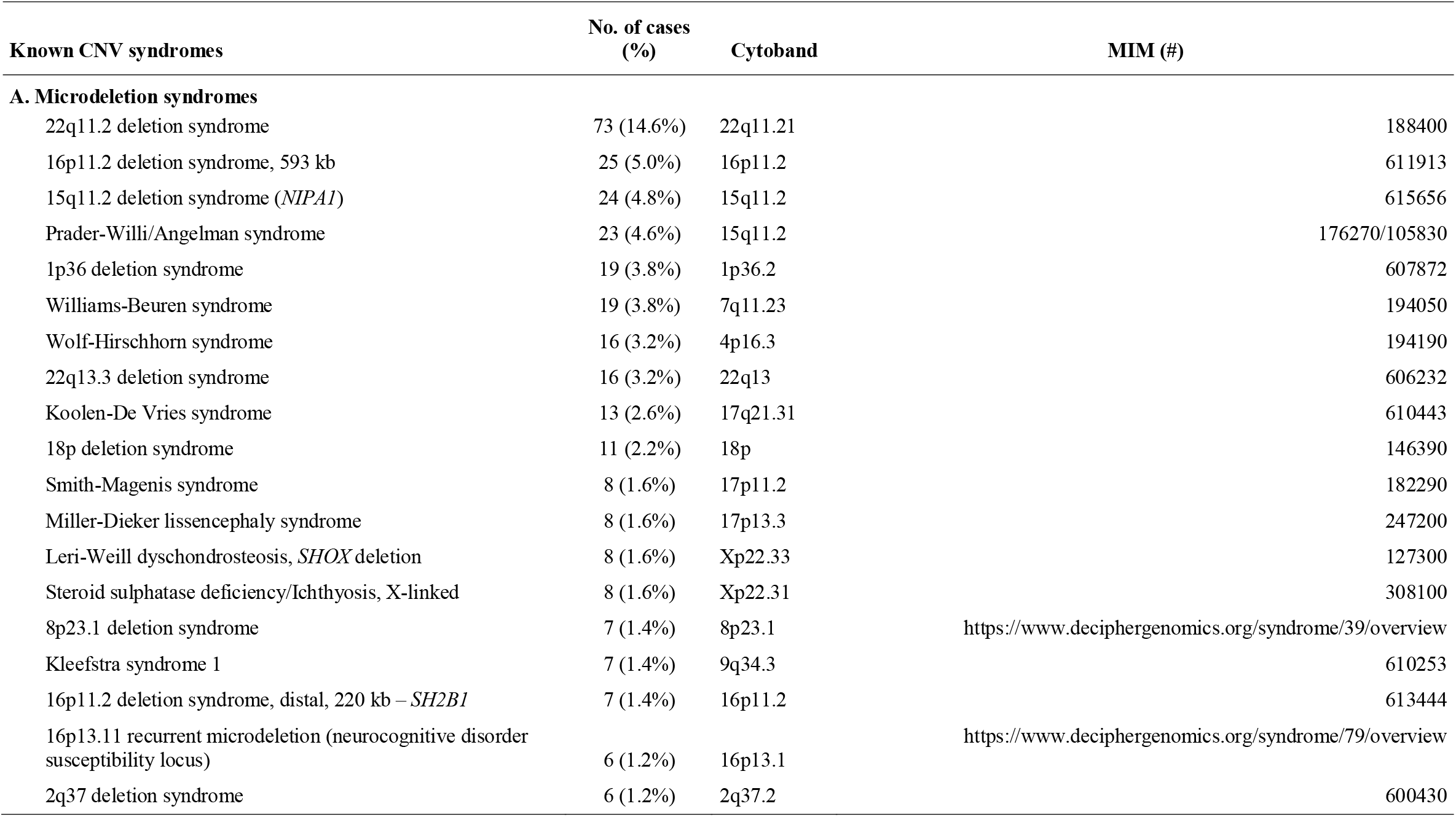
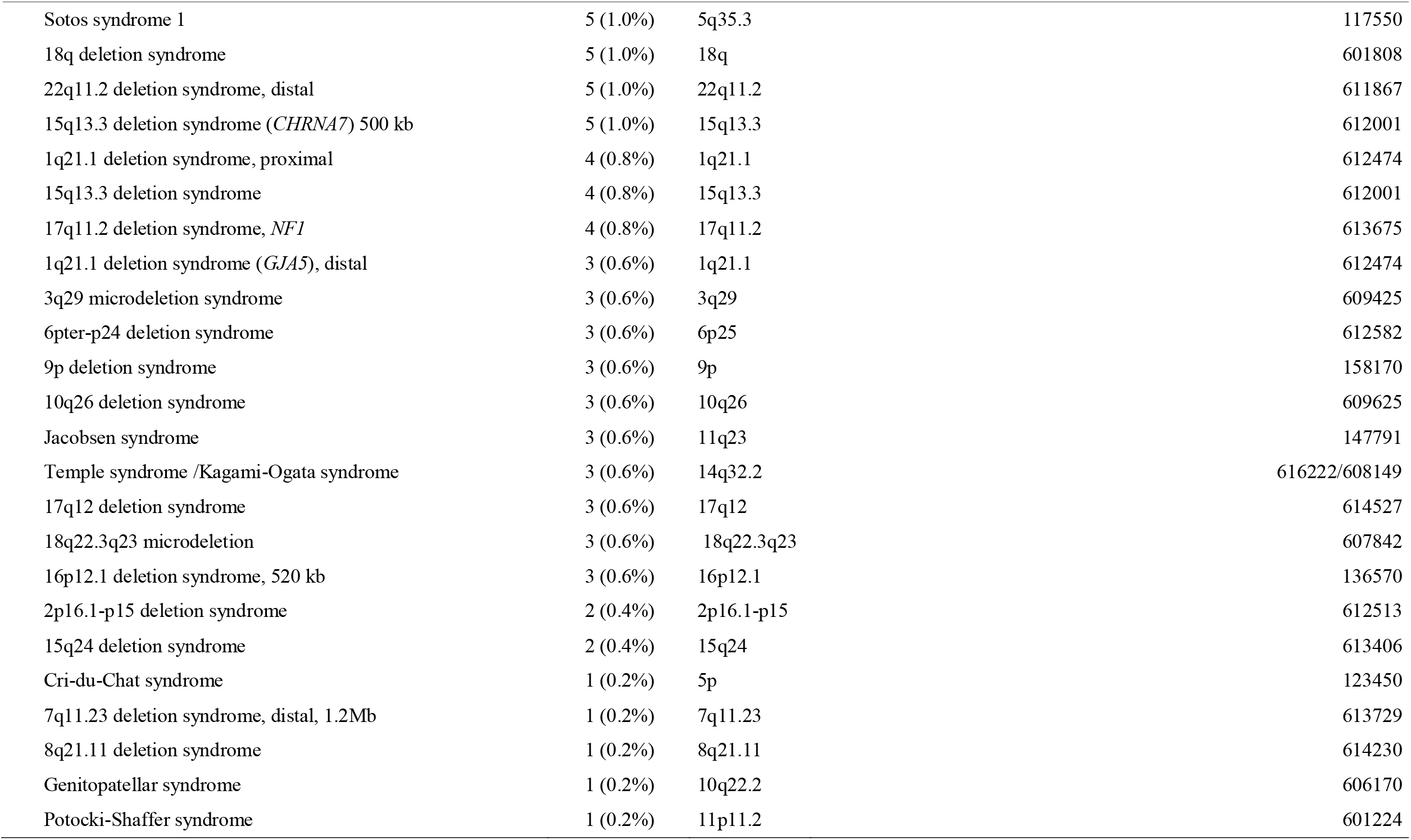
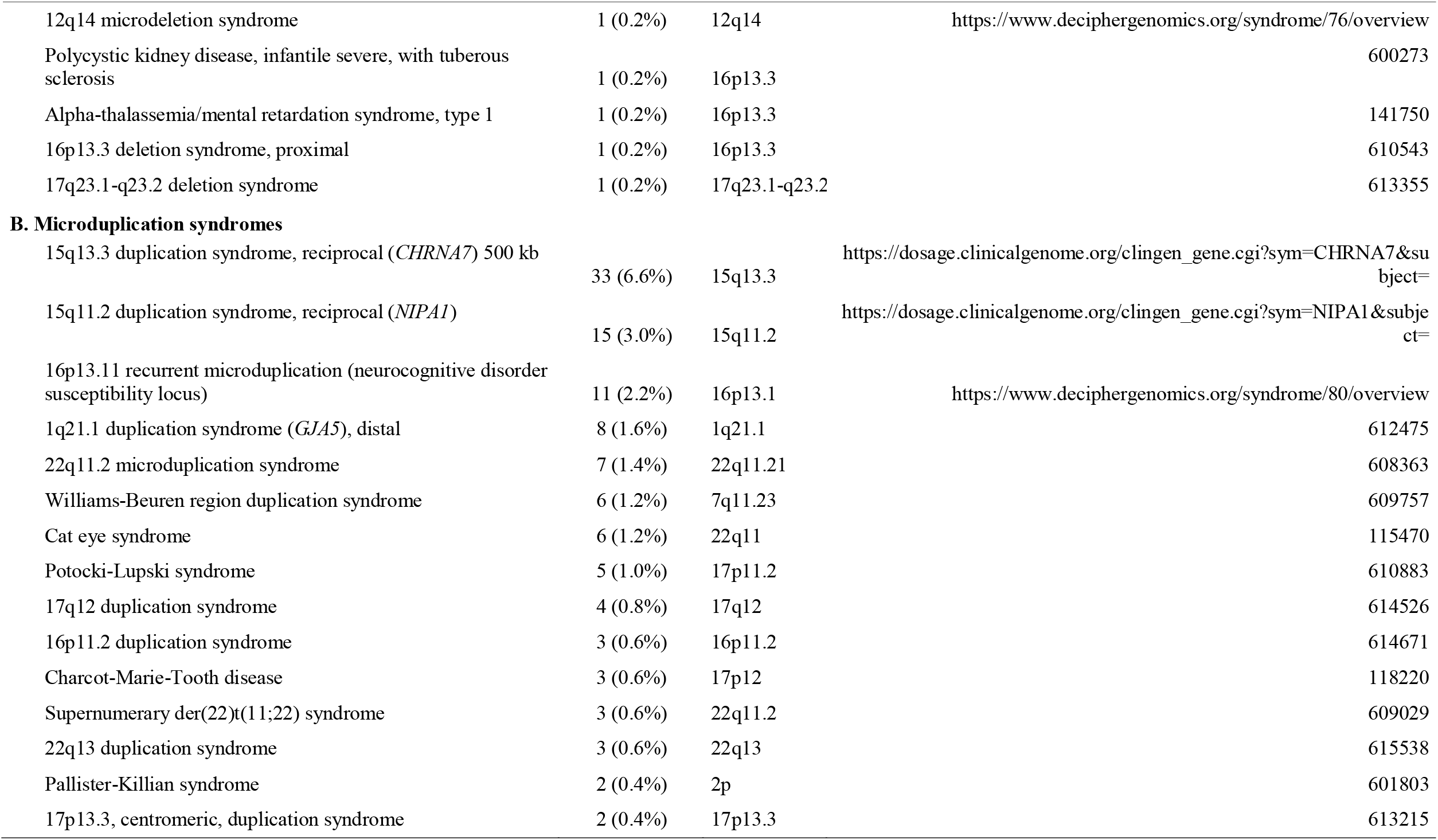
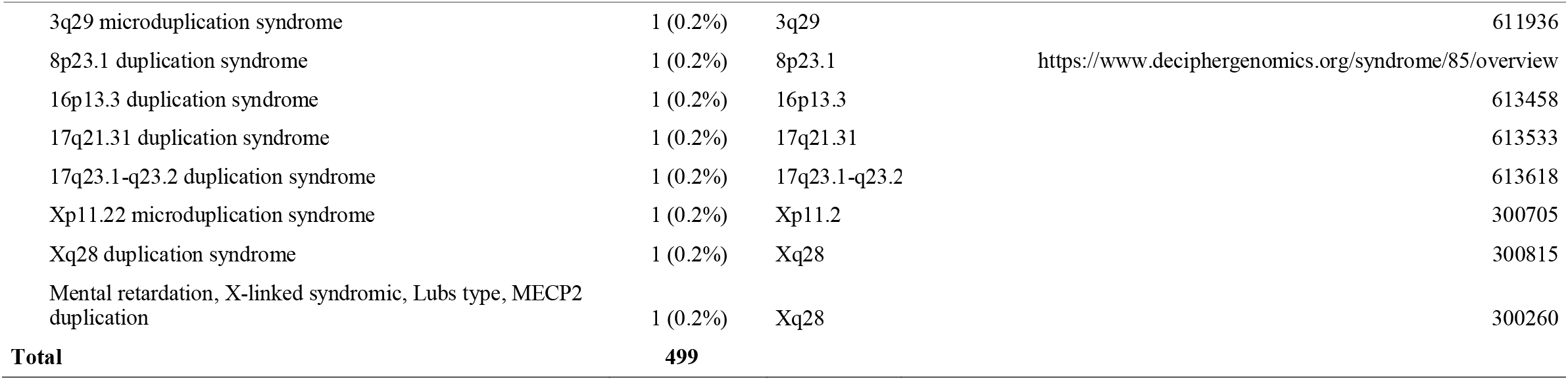
Frequency of the known copy number variation (CNV) syndromes

**Figure 4.**
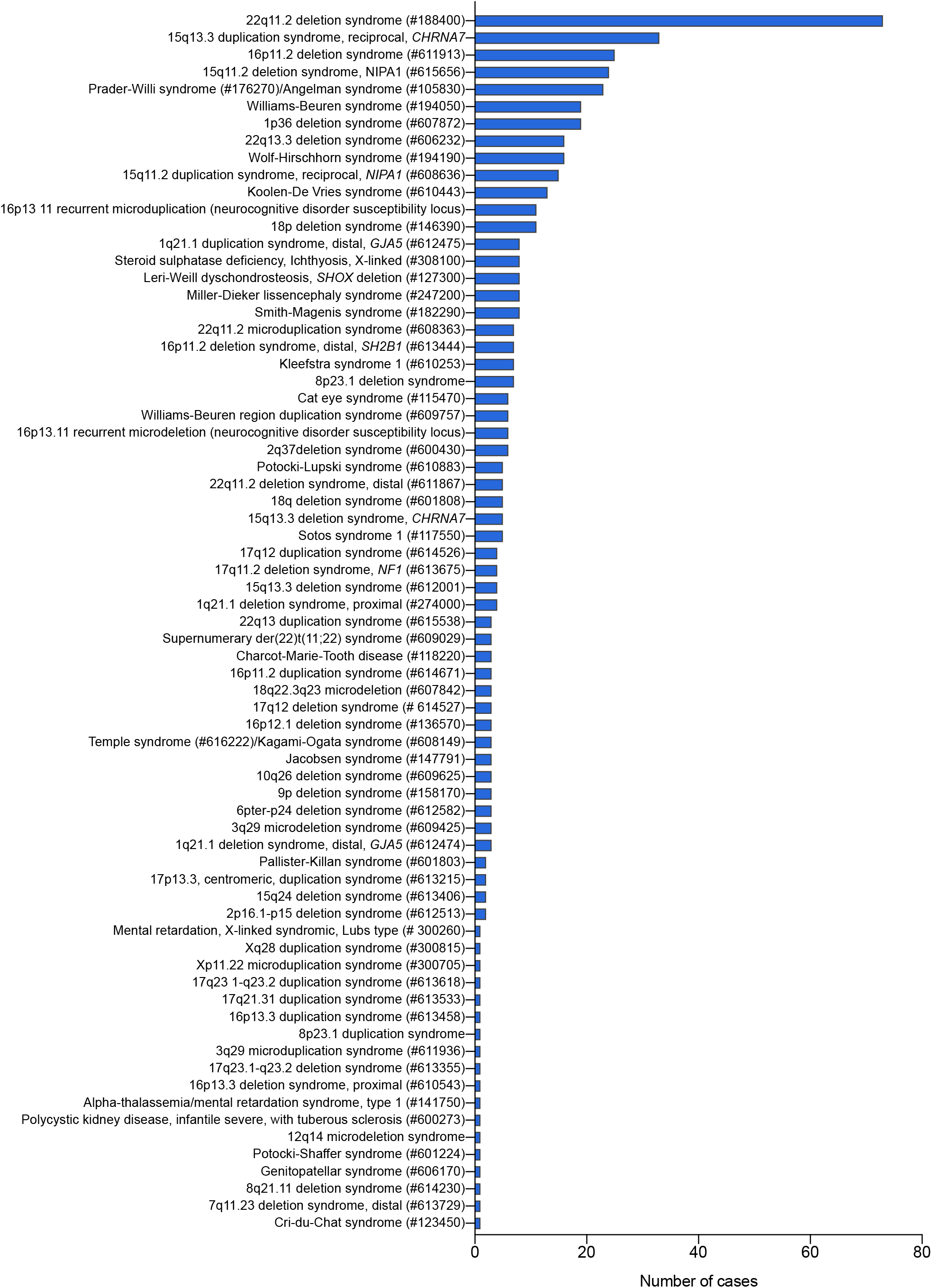
Distribution of microduplication and microdeletion syndromes. The histogram shows the frequency of microduplication and microdeletion syndromes identified in a total of 499 patients with neurodevelopmental disorders. Results are displayed in the descending order of frequency.

**Figure 5.**
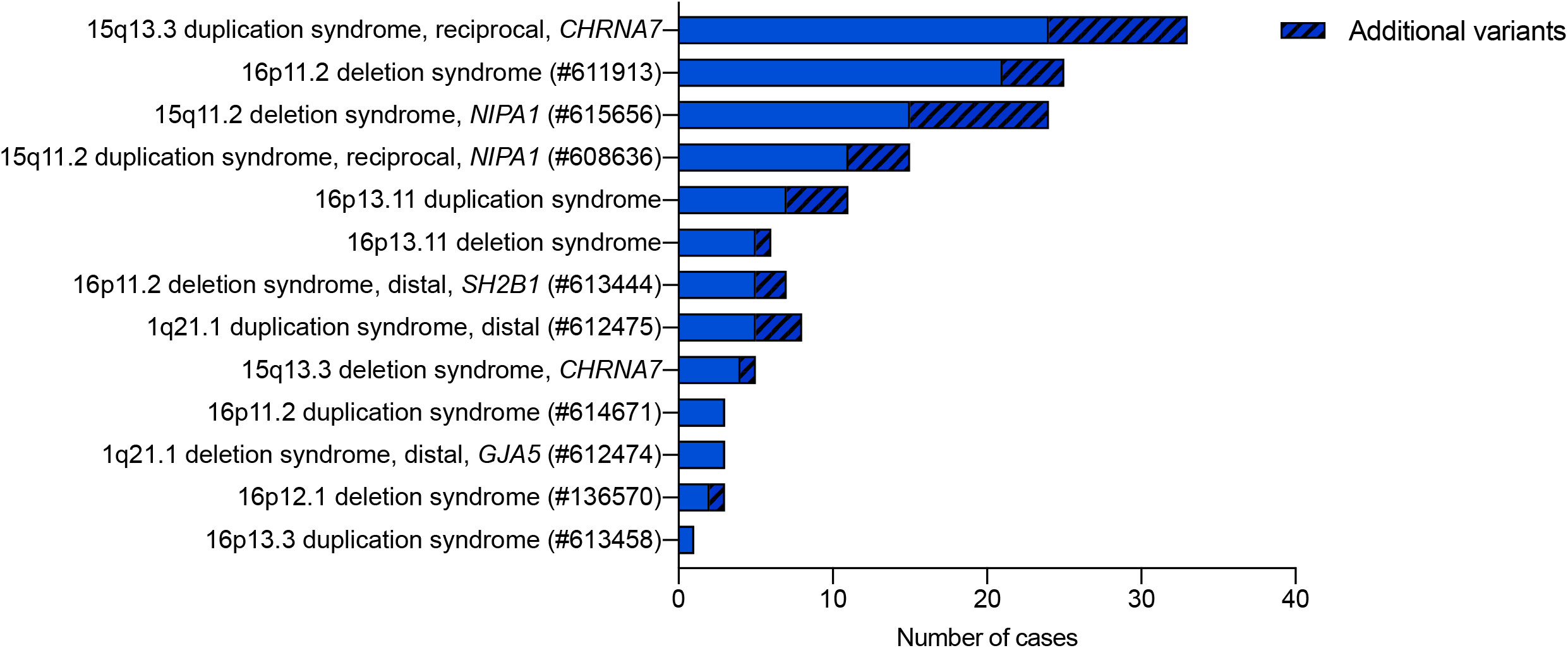
Frequency of syndromes with incomplete penetrance associated with predisposing CNVs, and additional variants. The histogram shows the frequency of a secondary CNV associated with a syndrome with incomplete penetrance.

Lastly, a total of 110 patients carried likely pathogenic CNVs, which represented only 5% of the alterations in our cohort (110/2,046). The third category of CNVs, the VUS accounted for 42% (868/2,046) of the cases. The description of all individual Pathogenic, Likely Pathogenic CNVs, and VUS can be found in **Supplementary Tables 1-3**.

### Copy neutral regions of homozygosity (ROH)

Copy neutral ROH were observed in 264 patients and were divided in two categories, according to the supposed origin of the ROH. Twenty-seven (10%) were classified as UPD, while only 15 were classified as Pathogenic variants (chromosomes 7, 11, 14 and 15). The most common UPD was UPD15 (8/27), followed by UPD14 (5/27) and UPD7 (4/27), each corresponding to a frequency of 33%, 19%, and 11%, respectively (**Figure 6**). In the remaining 237 individuals carrying ROHs, 90% of the cases, the presence of blocks of homozygosity was interpreted as indicative of identity by descent.

**Figure 6.**
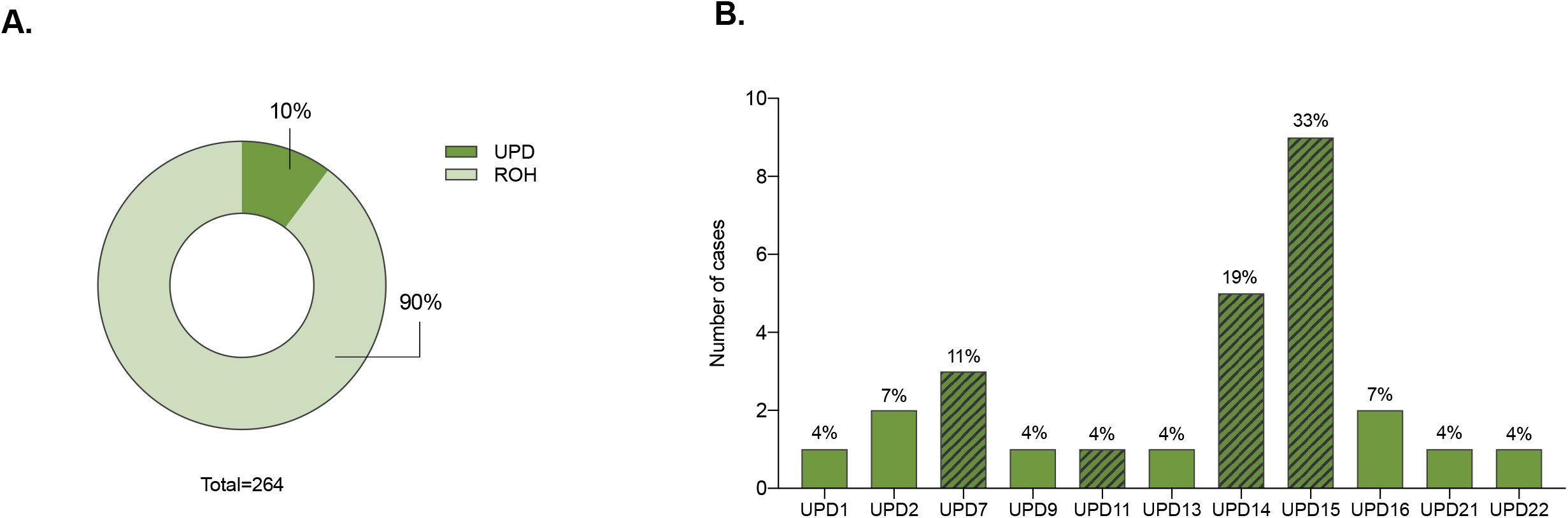
Frequency of copy neutral regions of homozygosity (ROH). (A) A total of 264 patients carried ROHs, 27 (10%) corresponding to uniparental disomy (UPD) cases. The remaining ROH in the other 237 patients (90%) were considered associated to different degrees of identity by descendent. (B) The histogram shows the frequency of UPDs per chromosome detected in our cohort. The crossed pattern represents pathogenic UPD known to encompass imprinting regions.

## DISCUSSION

In this study, we report the largest Brazilian cohort of patients investigated in by CMA. An overall diagnostic yield of 20% was achieved in our cohort, which is similar to what was found in other studies^8–11^. The copy number data presented here serve as a valuable clinical resource, and show once more the massive contribution of copy number changes in postnatal diagnosis.

CMA has been recommended and practiced routinely in USA and Europe as the first-tier test for patients with neurodevelopmental disorders and congenital abnormalities since 2010^1^. Nonetheless, in Brazil, the use of CMA is still limited due to its high cost. It is relevant to mention that the healthcare system in Brazil is a complex mixture of public and private funding, with governance and ownership arrangements. The public sector is one of the world’s largest single payer healthcare systems, but complementing this scenario there is a significant and large private sector with great investments; though, it is estimated that only ∼26% of Brazilians have a private health insurance, being the coverage mostly concentrated in the urban areas of the Southeastern part of the country^17^. Currently, nearly all patients referred for CMA come from the private sector, and even so the health insurances require that G-banded karyotype be used as the first genetic test; the patients with normal results being subsequently referred for investigation by microarrays. In contrast, in the public sector, CMA is not even offered to the patients; the price established by the government for the total genetic investigation of a patient do not pay even the costs of material for a single CMA. In practice, CMA is provided for few patients at Public Universities or Institutions, when it is linked to specific projects and research grants. However, this situation is not only a Brazilian peculiarity. In fact, even in countries with better economic conditions, private laboratories can contribute to a class disparity with regard to access to medical analyzes.

Although a large number of the patients in our cohort have been previously investigated by G-banded karyotype, we found 34 cases of aneuploidies, in which trisomy 21 correspond to the most frequent chromosomal disorder. The patients with Down syndrome were referred for CMA for presenting autistic features to search for other CNVs associated with autism spectrum disorders, but in none of the cases, additional CNVs were detected. It is noteworthy that the presence of autism spectrum disorders in individuals with Down syndrome has been well documented for several years^18,19^, therefore, the analysis through CMA appears to be a puzzling in economic terms.

The second most common aneuploidy detected was the Klinefelter syndrome; also in this case, behavioral disorders were the main reason for CMA referral. Epidemiological studies of autism have demonstrated that in about 10-25% of cases there is a correlation with other known medical conditions, often chromosome alterations^20^. In fact, autistic features may be more common in individuals with sex chromosome aneuploidies than generally believed^21^.

Marker chromosomes from both autosomes and sex chromosomes represented 3% of the diagnostic alterations. Except for the X and Y chromosomes, all were supernumerary. The correlation of specific supernumerary marker chromosomes (SMC) with distinct clinical features have been demonstrated for some syndromes, for example: i(18p) syndrome (MIM#146390), i(12p)-(Pallister-Killian) syndrome (MIM#601803), der(22) and Cat eye syndromes (MIM#609029 and MIM#115470, respectively)^22^. The only marker that represents a derivative chromosome in our dataset characterize the Emanuel syndrome (MIM#609029), and results from missegregation of the only known recurrent, non-Robertsonian, constitutional translocation in humans [der(22)t(11;22)(q23;q11.2)]. All these disorders associated with SMC were identified in our cohort. Although it was not possible to obtain cytogenetic characterization of all markers, it is known that an inverted duplicated chromosome 15^23^ is the most common of the heterogeneous group that constitute the supernumerary marker chromosomes.

The microdeletion and microduplication syndromes represented the largest proportion of pathogenic variants in our cohort (25%). Many of them harbor genomic hotspots flanked by homologous segmental duplications prone to unequal crossing over, and have high elevated *de novo* mutation rates, generally with similar CNV sizes^24,25^. In this study, we detected 71 distinct microdeletion/microduplication syndromes in a total of 499 individuals, in which deletions were twice as common than duplications (*n*= 48 deletions vs *n*=23 duplications). Among the five most frequent syndromes, shown in **Figure 4**, four have incomplete penetrance and variable expressivity: 22q11.2 deletion (MIM#188400), 15q13.3 duplication^16^, 16p11.2 deletion (MIM#611913) and 15q11.2 deletion (MIM#615656). Such susceptibility CNVs impose a challenge in genetic counseling since they are present in the normal population but enriched in individuals with various neurodevelopmental disorders. Moreover, these CNVs are often inherited from a normal or mildly affected parent and they lack phenotypic specificity, being associated with a variety of neuropsychiatric disorders, medical conditions, and variable dysmorphisms.

The 22q11.2 deletion was the most frequent syndrome in our cohort, reflecting the same frequency reported by other large population studies^26^. In particular, the 15q13.3 duplication encompassing the *CHRNA7* gene has been consistently associated to several neurodevelopmental disorders^27–29^. However, overlapping duplications in this genomic region were also documented in many individuals of the general population (∼0.6% - estimated prevalence of 1:174-186 individuals^30^) and, in almost all cases investigated, patients inherited the duplication from clinically normal parents. The high frequency of duplications of this segment in the general population, together with the lack of enrichment in clinical cohorts^31,32^, indicate that, if this variant has any clinical impact, the penetrance would be very low. Indeed, this variant was historically classified as pathogenic, and the data presented reflects a retrospective analysis; currently, 15q13.3 duplications have been classified as probably benign in light of recent evidence, and in accordance to databases such as DECIPHER and ClinGen.

The recurrent 15q11.2 deletion (BP1-BP2), which includes the *CYFIP1, NIPA1, NIPA2, TUBGCP5* genes, is consistently associated with neurocognitive function. Jonch et al. (2019) performed a comprehensive meta-analysis on new and previously published individuals with 15q11.2 deletions, comparing data across 20 studies. The case-control study using their clinical cohort compared to controls in the UK Biobank cohort showed enrichment of the deletion in the patient population. Nonetheless, the reciprocal duplication of the 15q11.2 region has refuted clinical significance^31,33^. Duplications of this region are common in the general population and the majority of case-control studies have observed a lack of enrichment in the clinical population. Recent studies of duplication carriers identified through cohort studies in the general population have also shown that carrier individuals perform similarly to non-carrier controls on neurocognitive tests. Therefore, the triplosensitivity is considered unlikely dosage sensitivity.

The estimated frequency of the 16p11.2 deletion syndrome is about 1–5/10,000 in the general population. Research based on the ClinGen database suggests that 16p11.2 deletions are the second most common microdeletion, occurring in one of every 235 individuals tested with intellectual and developmental disabilities. Interestingly, this deletion was identified in nearly 1% of individuals with autism and has been found in up to 0.001% of those with psychiatric disorders^34–36^. Nonetheless, the phenotypic spectrum associated with this deletion is much wider and includes delays in speech or motor development, language impairment, low muscle tone, hypo-or hyperreflexia, a tendency towards obesity, short stature, and several facial dysmorphisms. The syndrome classically involves a heterozygous microdeletion of ∼600 kb, containing 29 protein coding genes; although the majority of cases reported are *de novo*, the deletion is inherited in an autosomal dominant pattern in 20% of the times; an equal sex ratio has been reported^37^.

Considering all evidence from association studies about the susceptibility CNVs for neurodevelopmental disorders, the general consensus is that there must be additional modifiers that influence the expression of these variants. A “two-hit”, or second site, model has been suggested for several syndromes^38,39^. Notably, the vast majority of the syndromes with incomplete penetrance, as shown in **Figure 5** from our cohort, present an additional CNV in part of the patients, mostly classified as VUS. Girirajan et al. (2010) demonstrated that affected individuals with a microdeletion on chromosome 16p12.1 were more likely to have additional large copy-number variants than control individuals^39^. This data supported an oligogenic basis, in which the compound effect of a relatively small number of rare variants of large effect contributes to the heterogeneity of genomic disorders. The authors also identified other known genomic disorders, each defined by a specific CNV, in which the affected children were more likely to carry multiple copy number variants than controls^38^. However, in most cases, the supposed second hit is not identified; it can be speculated that single nucleotide variant(s) could contribute, but there are not data supporting this claim. It is relevant to mention that investigation of the parents is mandatory in cases of those susceptibility CNVs for proper genetic counselling^15^. However, in our study we were unable to obtain information about inheritance pattern for all cases, because the patients and parents were not necessarily investigated at the same centers or in the same period of time.

In cases where the patient present one or more VUS, the recommendation is to investigate the parents to determine whether the CNV has been inherited or represents a *de novo* mutation^15^. While the latter may lead to a reclassification of the VUS as pathogenic or likely pathogenic, the inherited variants remain classified as VUS. In our cohort, we have information on segregation in a minority of the cases: among 126 patients who presented an autosomal VUS and segregation was tested, we found that 125 of the variants where inherited. It is important to remember that, regardless of being inherited, these VUS may still contribute to the patients’ phenotype, and must be reported. While it is undeniable that the resulting 0.8% *de novo* alterations have some impact in diagnosis and genetic counselling, the healthcare context should be considered in the decision to test the parents. In the present cohort, we performed 252 CMA tests in parents, but were able to reclassify the VUS in a single case. For all remaining 125 cases, the parental tests did not add any useful information. When resources are limited, such as in Brazil and in many other countries, we obtain a better cost-benefit testing other 252 patients instead of investigating VUS segregation. Therefore, we would not recommend testing segregation of VUS using CMA in the public healthcare in Brazil or in other developing countries in the current situation. Nonetheless, segregation analysis can be performed with cheaper techniques such as real time PCR; when the VUS is *de novo* it is necessary to consider whether it is itself causative of the clinical condition due to the consequent gene imbalance or whether it alters the expression of contiguous genes, in any case in a condition of dominance. In contrast, if the VUS is inherited from a healthy parent, it cannot be excluded that, together with an allelic variant in the homologous chromosome, the VUS is at the basis of the clinical condition of the patient, as a recessive condition. In this second case, the knowledge of the risk of recurrence of the disease in subsequent pregnancies becomes fundamental.

Importantly, with the incorporation of many robust SNP-array platforms in the clinical routine, many studies have identified large ROH in patients with a wide variety of clinical features^40–43^. Depending on chromosomal distribution and cumulative extent, it may indicate either UPD or parental consanguinity^44^. When ROH >10 Mb are detected in a single chromosome subject to imprinting, the first possibility to be considered is UPD, an event that arises as a consequence of the correction of a meiotic or early mitotic error. In such cases, the presence of two copies of the same chromosome inherited from only of the parents is considered as pathogenic *per se*. However, UPD in other chromosomes, besides increasing the probability of deleterious mutations in homozygosity^45^, could be originated from a trisomy rescue, which may have further implications, such as a trisomic cell line in an tissue other than the investigated one or during development. Contrarily, the presence of many ROH throughout the genome is an indication of consanguinity, and the chance of inheritance of recessive monogenic disorders increases with the degree of relatedness. It has been demonstrated that the occurrence of multiple congenital anomalies and other significant clinical problems is higher among children of first cousins (4.4%) and second cousins (3.6%), compared to unrelated parents^46^. The rate of consanguinity in the different regions of Brazil is expected to be very heterogeneous and estimates are scarce. A recent paper indicates that some degree of inbreeding may be present in 26,5% of patients with developmental disorders of the South of Brazil^2^. However, when clinically more relevant kinship of 1^st^ to 5^th^ degree is considered, they find consanguinity in ∼ 8.5% of the cases. In our cohort, 237 out of the 264 cases of ROH were interpreted as the result of consanguinity, reflecting a frequency of 4,7% (237/5,778), which is not much different from the 6% reported by Wang et al. (2015) in the North American population, using a cutoff similar to ours (minimum 10 Mb of ROH)^44^.

In summary, we reported copy number data from patients with neurodevelopmental disorders and congenital anomalies in the largest Brazilian cohort investigated by CMA reported so far. Also, considering that the results are originated from SNP microarrays, it can be used for comparison and investigation of the genomic etiology across multiple disorders in other studies.

## Data Availability

All data generated or analysed during this study are included in this published article [and its supplementary information files].

## ACKNOWLEDGMENTS

This work was supported by FAPESP grant (2013/08028-1), and FAPESP Innovative Research in Small Business (12/50981-5). D.V. was supported by a FAPESP scholarship (2014/17132-0).

